# Artificial Intelligence in Gastrointestinal Endoscopy: A Comprehensive Systematic Review

**DOI:** 10.1101/2025.07.02.25330317

**Authors:** Rajvi Chaudhary, Pathik Dhangar, Aditya Chaudhary

**Affiliations:** B. J Medical College, Ahmedabad, Gujarat 380016; Department of Internal Medicine, All India Institute of Medical Sciences, Rishikesh, Uttarakhand 249203, India

**Author notes:** **Corresponding author Pathik Dhangar**, Department of Internal Medicine, Sixth Floor, College Block, All India Institute of Medical Sciences (AIIMS), Rishikesh, Uttarakhand 249203, India. **Contributors** Chaudhary R and Chaudhary A contributed to the data collection and data analysis and was involved in manuscript writing. Dhangar P reviewed the draft. **Data sharing** It will be made available to others upon request to the corresponding author. **Funding source** None.

## Abstract

Artificial intelligence (AI) has emerged as a transformative force in gastrointestinal (GI) endoscopy, offering enhancements in diagnostic accuracy, lesion detection, and procedural efficiency. This systematic review consolidates current evidence from original research articles on AI applications across various endoscopic procedures, including colonoscopy, esophagogastroduodenoscopy (EGD), and capsule endoscopy. The review highlights AI’s performance in detecting colorectal polyps, gastric cancer, small bowel lesions, and inflammatory bowel disease (IBD). While AI demonstrates high sensitivity and specificity across multiple studies, challenges such as data heterogeneity, integration into clinical workflows, and the need for external validation persist. Future research should focus on large-scale, multicenter trials to validate AI’s efficacy in diverse clinical settings.

## Introduction

Gastrointestinal endoscopy is pivotal in diagnosing and managing GI diseases. However, its effectiveness is often limited by operator dependency and variability in lesion detection. Artificial intelligence (AI), particularly machine learning and deep learning algorithms, offers the potential to augment endoscopic procedures by providing real-time analysis and decision support. This review aims to systematically evaluate the current landscape of AI applications in endoscopy, assessing diagnostic performance, clinical utility, and areas requiring further investigation.

## Methods

### Eligibility Criteria

We included original peer-reviewed studies evaluating the application of artificial intelligence (AI), including machine learning (ML) and deep learning (DL), in gastrointestinal (GI) endoscopy. Eligible studies had to report quantitative outcomes such as sensitivity, specificity, diagnostic accuracy, or area under the receiver operating characteristic curve (AUC).

Exclusion criteria included:

- Non-original research (e.g., reviews, editorials)
- Conference abstracts without full texts
- Case reports and small series (<10 patients)
- Studies not using AI/ML algorithms
- Articles not in English

### Information Sources

We systematically searched the following databases from inception to May 31, 2025:

- PubMed/MEDLINE
- Scopus
- Embase
- Web of Science
- Cochrane Library

In addition, we screened the references of included articles for potentially eligible studies.

### Search Strategy

The PubMed search strategy included the following terms:

(“Artificial Intelligence” OR “Machine Learning” OR “Deep Learning” OR “Neural Networks”) AND

(“Endoscopy” OR “Colonoscopy” OR “Capsule Endoscopy” OR “Gastroscopy” OR “Enteroscopy”) AND

(“Gastrointestinal” OR “GI” OR “Ulcerative Colitis” OR “Crohn’s Disease” OR “Colorectal Cancer” OR “Polyps” OR “Gastric Cancer”)

Similar strategies were adapted for other databases using controlled vocabulary (e.g., MeSH and Emtree terms) where applicable.

### Study Selection

Two independent reviewers (Reviewer A and B) screened titles and abstracts. Full-text articles were retrieved for studies meeting inclusion criteria or if eligibility was unclear. Discrepancies were resolved by discussion or a third reviewer.

### Risk of Bias Assessment

For diagnostic accuracy studies, we used the QUADAS-2 tool to assess the risk of bias. Each study was evaluated in four domains: patient selection, index test, reference standard, and flow/timing. Disagreements were resolved through discussion.

### Data Synthesis

We performed a qualitative synthesis due to methodological heterogeneity. Studies were grouped by:

- Procedure type: colonoscopy, capsule endoscopy, upper GI endoscopy
- AI function: detection, classification, disease activity scoring
- Clinical setting: screening vs. diagnostic

## Results

### AI in Colonoscopy

Colonoscopy remains the gold standard for colorectal cancer (CRC) screening. However, adenoma miss rates can be significant, leading to interval cancers. AI-assisted colonoscopy, particularly computer-aided detection (CADe) systems, has shown significant improvements in adenoma detection rates (ADR).

#### Colorectal Cancer

##### Real-time Polyp Detection

A study by Schauer et al. demonstrated that a real-time automatic polyp detection system improved the polyp detection rate (PDR) in a clinical environment.[1]. AI in colonoscopy increases the detection of inconspicuous polyps more commonly missed by endoscopist. It can also elevate accuracy of polyp characterization when used by regular endoscopist to nearly that of highly trained expert interventionalist. [2]

##### Multicenter Randomized Controlled Trials

Luo et al. conducted a multicenter RCT showing that AI-assisted colonoscopy improved overall ADR, advanced ADR, and ADR of both expert and non-expert attending endoscopists.[3]

##### Effect on Novice Endoscopists

A study by Yao et al. compared lesion detection capabilities among novices, AI-assisted novices, and experts, finding that AI assistance significantly improved the performance of novice endoscopists.[4]

##### Cost-Effectiveness

Areia et al. analyzed the cost-effectiveness of AI in screening colonoscopy, concluding that AI detection tools are a cost-saving strategy to further prevent CRC incidence and mortality.[5]

#### Inflammatory bowel disease

AI has been utilized to assess endoscopic activity in IBD, aiming to reduce inter-observer variability and improve disease monitoring.

##### Ulcerative Colitis Assessment

Lacucci et al. developed a deep learning model that accurately assessed endoscopic disease activity in ulcerative colitis, correlating well with histological findings. [6]

### AI in Capsule Endoscopy

Capsule endoscopy (CE) is a non-invasive method for visualizing the small intestine but is time-consuming and subject to human error. AI applications have been developed to address these challenges.

#### Diagnostic Yield

A study by Spada et al. assessed the non-inferiority of AI-assisted reading versus standard reading in detecting small bowel bleeding lesions, finding that AI-assisted reading was non-inferior and significantly reduced reading time. [7]

#### Real-World Application

O’Hara et al. evaluated an AI-enabled small bowel capsule in a real-world setting, demonstrating its effectiveness in reducing reading time without compromising diagnostic accuracy. [8]

#### Pan-Enteric Capsule Endoscopy

A study by Brodersen et al. examined AI-assisted analysis of pan-enteric capsule endoscopy in patients suspected of Crohn’s disease, showing high agreement between AI-assisted assessments and standard evaluations.[9]

### AI in Upper GI Endoscopy

AI applications in upper GI endoscopy focus on detecting conditions such as gastric cancer and Helicobacter pylori infection.

#### Gastric Cancer Detection

Niikura et al. developed a convolutional neural network (CNN) system that achieved high sensitivity in detecting gastric cancer from endoscopic images. [10]

#### Helicobacter pylori Diagnosis

Parkash et al. created an AI system that accurately diagnosed H. pylori infection using endoscopic images, outperforming experienced endoscopists. [11]

## Review

### Enhancing Diagnostic Accuracy and Efficiency

Artificial intelligence (AI) has demonstrated significant potential in augmenting the diagnostic capabilities of gastrointestinal (GI) endoscopy. In colonoscopy, AI-assisted systems have consistently improved adenoma detection rates (ADR), a key quality indicator. For instance, a multicenter randomized controlled trial (RCT) reported that AI-assisted colonoscopy significantly increased ADR compared to standard colonoscopy, irrespective of endoscopist experience, system type, or healthcare setting.[12] Furthermore, AI has been instrumental in reducing the miss rate of colorectal neoplasia, with studies indicating approximately a two-fold reduction, thereby supporting its benefit in minimizing perceptual errors for small and subtle lesions.[13]

In capsule endoscopy, AI applications have been developed to address challenges such as time-consuming procedures and human error. A study assessing AI-assisted capsule endoscopy reading in suspected small bowel bleeding found that AI-assisted reading provided more accurate and faster detection of clinically relevant lesions than standard reading.[7] Similarly, real-world usage of a validated AI model for capsule image review demonstrated its effectiveness in reducing reading time without compromising diagnostic accuracy. [7]

In upper GI endoscopy, AI has been employed to detect conditions such as gastric cancer and Helicobacter pylori infection. A study evaluating the diagnostic ability of AI in detecting early upper gastrointestinal cancer (EUGIC) using endoscopic images found that AI achieved high diagnostic accuracy, with sensitivity like that of expert endoscopists. [14][15]. Additionally, AI has shown high diagnostic accuracy for detecting H. pylori infection using endoscopic images.[11]

In the context of inflammatory bowel disease (IBD), AI has been utilized to assess endoscopic activity, aiming to reduce inter-observer variability and improve disease monitoring. Studies have indicated that AI-assisted endoscopy in IBD is a rapidly growing research field, with promising technical results in automated endoscopic scoring and real-time prediction of histological disease. [16] Moreover, integrating AI into IBD management has the potential to revolutionize clinical practice by providing accurate assessments of endoscopy and histology, offering precise evaluations of disease activity, standardized scoring, and outcome prediction.[17]

### AI in Colonoscopy: Enhancing Detection Rates

Artificial intelligence (AI) has significantly improved adenoma detection rates (ADR) during colonoscopy.[18] A comprehensive meta-analysis of 28 randomized controlled trials (RCTs) involving 23,861 participants demonstrated a 20% increase in ADR (risk ratio [RR], 1.20; 95% confidence interval [CI], 1.14–1.27; P < .01) and a 55% decrease in adenoma miss rate (RR, 0.45; 95% CI, 0.37– 0.54; P < .01) with AI-assisted colonoscopy. These improvements were consistent across various subgroups, including expert endoscopists and different healthcare settings.[12]

Further analysis revealed that AI-assisted colonoscopy significantly increased the detection of diminutive lesions, contributing to the overall improvement in ADR. However, there was no significant difference in the detection of advanced adenomas or sessile serrated lesions.[12] Additionally, AI-assisted procedures showed a slight increase in withdrawal time (mean difference of 0.15 minutes) and a 39% increase in non-neoplastic resection rates, indicating a need for careful consideration of potential overdiagnosis.[19]

### AI in Capsule Endoscopy: Streamlining Small Bowel Evaluation

Capsule endoscopy (CE) is a valuable tool for evaluating small bowel pathology, but it is time-consuming and subject to human error. A multicenter prospective study assessed the efficacy of AI-assisted reading in patients with suspected small bowel bleeding. The study found that AI-assisted reading had a higher diagnostic yield (73.7%) compared to standard reading (62.4%) and significantly reduced the mean reading time from 33.7 minutes to 3.8 minutes.[7]

Moreover, a convolutional neural network developed for detecting blood and hematic residues in the small bowel lumen achieved an accuracy of 98.5% and a precision of 98.7%, demonstrating the potential of AI to enhance the diagnostic accuracy of CE. [20]

### AI in Upper GI Endoscopy: Advancements and Challenges

AI applications in upper gastrointestinal (GI) endoscopy have shown promise in detecting neoplasia. Studies have reported that computer-aided detection (CADe) systems achieved sensitivities ranging from 83% to 93% in identifying esophageal squamous cell neoplasia, Barrett’s esophagus-related neoplasia, and gastric cancer.[21] However, the integration of AI into clinical practice faces challenges, including the need for large, well-annotated datasets and addressing potential biases in training algorithms.[22]

### AI in Inflammatory Bowel Disease: Towards Standardized Assessment

In inflammatory bowel disease (IBD), AI has been utilized to assess endoscopic disease activity, aiming to reduce inter-observer variability. A systematic review found that AI models assessing ulcerative colitis (UC) endoscopic activity achieved accuracies ranging from 86.5% to 94.5%.

Additionally, AI-assisted capsule endoscopy reading significantly reduced reading time while maintaining high accuracy. AI models have also demonstrated the ability to differentiate between IBD and non-IBD, UC and non-UC, and UC and Crohn’s disease with high accuracy.[23]

Furthermore, a meta-analysis evaluating AI’s ability to diagnose endoscopic remission in UC reported a combined sensitivity of 87% and specificity of 92%, with an area under the curve (AUC) of 0.96, indicating excellent diagnostic performance.[24]

### Challenges and Future Directions

Despite the promising advancements, several challenges hinder the widespread clinical implementation of AI in GI endoscopy. One significant concern is the generalizability of AI models, as many studies are based on retrospective reviews of selected images, necessitating further validation in prospective trials.[25] Additionally, endoscopists’ knowledge, perceptions, and attitudes toward the use of AI in endoscopy vary, highlighting the need for comprehensive education and training to facilitate adoption.[26]

Furthermore, the integration of AI into clinical workflows requires addressing technical, ethical, and regulatory considerations. Establishing key research questions for the implementation of AI in colonoscopy is crucial to guide future studies and ensure that AI applications align with clinical needs.[27] Moreover, the development of AI algorithms for endoscopy quality assessment, including monitoring of blind spots, bowel preparation, and withdrawal time, can enhance the overall quality of endoscopic procedures.[28]

## Conclusion

Artificial intelligence (AI) is rapidly transforming the landscape of gastrointestinal (GI) endoscopy, offering unprecedented improvements in lesion detection, characterization, workflow efficiency, and real-time decision-making. Robust evidence from randomized controlled trials and prospective studies demonstrates that AI-enhanced colonoscopy significantly increases adenoma detection rates while reducing miss rates, especially for diminutive lesions. In capsule endoscopy, AI markedly shortens reading times without compromising diagnostic accuracy, thereby addressing one of its major clinical limitations.

Furthermore, AI applications in upper GI endoscopy show promising sensitivity in identifying early neoplastic lesions, although generalizability and validation remain critical hurdles. In inflammatory bowel disease, AI models have demonstrated excellent correlation with histologic disease activity and offer potential for standardized, reproducible scoring. Despite these advances, challenges persist, including data heterogeneity, limited external validation, and variability in clinician acceptance.

Moving forward, the focus must shift toward multicenter validation, integration of AI tools into clinical workflows, and regulatory clarity to ensure safety, equity, and effectiveness. As AI continues to evolve, its successful implementation will depend not only on technological refinement but also on a multidisciplinary approach involving clinicians, data scientists, and regulatory bodies. AI in endoscopy is no longer a futuristic concept but an imminent clinical reality poised to redefine diagnostic accuracy, procedural quality, and patient outcomes in gastroenterology.

## Data Availability

All data produced are available online

